# Impact of conditional mobile data credits on caregivers’ use of an early childhood digital health intervention: a nested cluster-randomized controlled trial in Andean Peru

**DOI:** 10.64898/2026.10.07.26364726

**Authors:** Sarah Farnsworth Hatch, Leonel Aguilar, Dana McCoy, Günther Fink, Kristen Hinckley, Andreana Castellanos, Milagros Alvarado, Maria Catalina Gastiaburu Cabello, Stella Hartinger, Lena Jäggi

## Abstract

As internet and mobile phone access increases, digital health interventions are increasingly used to support remote and low-resource populations. In many settings, the health impact of digital interventions is constrained by limited or short-lived engagement. We tested if conditional mobile data credit incentives increase digital intervention engagement within a digital intervention trial. This nested cluster randomized controlled trial was conducted with caregivers of children 3-9 months at baseline. Of 72 neighborhood clusters from the treatment arm of the parent trial (evaluating a digital early childhood development intervention), 20 were randomly assigned to treatment and 52 as controls. Eligible participants had a phone provider through which mobile data credits could be most affordably administered. Treated participants received around 250 MB mobile data credits every two weeks, conditional upon using the digital intervention at least once in the same period. Credits were digitally administered via a third-party service. Digital intervention engagement (primary outcome) was monitored continuously until endline data collection approximately 2 years after assignment. Secondary outcomes included caregivers’ views, behaviors, and early childhood development. We used intent-to-treat analyses to assess all outcomes. This trial was registered as NCT05202106 at clinicaltrials.gov and was completed in 2025. Participants were recruited between September 2021 and March 2023 (N Treatment = 147; N Control = 333). Conditional mobile data credits extended engagement with the digital early childhood development intervention by 6.50 weeks on average (treatment mean = 26.19 weeks, control mean = 19.69; *p*=0.011). No significant impacts were identified on secondary outcomes. As such, mobile data credits may be a cost-effective way for increasing caregiver engagement with digital health intervention in low resource settings.

## Introduction

By 2022, 81.4% of 5.4 billion mobile phone subscribers had access to mobile internet, which coincides with rising literacy levels across Low– and Middle-Income Countries (LMIC).^1,2^ These converging factors point to the promise of mobile phones to provide preventive guidance, health promotion, and screening reminders to remote or low-resource populations who face reduced access to health resources.^3,4^

Digital early childhood development (ECD) interventions are one example of prevention platforms that provide low-intensity guidance to support child health and wellbeing at scale. By providing information to caregivers on how to promote children’s positive development from the earliest years, such interventions can support a strong foundation for cognitive, psychosocial, and physical growth throughout the life course.^5,6^ Artificial Intelligence– (AI-) may further increase the perceived relevance and usefulness of such platforms by enabling more personalized and responsive intervention elements.^7^ Some AI-supported digital ECD interventions have been found to promote caregiver and child outcomes in LMICs.^8,9^

Although self-guided digital interventions hold promise, low user engagement may undermine the potential benefits for families. Across diverse socioeconomic contexts, digital ECD program completion rates as low as 7 – 15% have been reported.^5^ Low engagement could be the result of difficulties with installation and use^10,11^ or decreases in caregivers’ perceived need for the intervention.^12^ In other digital health interventions in LMICs, users also report that major barriers to engagement include free time to engage in the program and operational constraints such as low phone storage or limited data.^5,13–15^

Operational barriers to engagement may be especially pronounced for families with limited financial resources. Families with constrained resources in LMICs may not be able to afford to purchase mobile data for an entire month, instead purchasing data packages for a fixed number of mega– or gigabytes. With these constraints, individuals may face limited internet access by complete disconnection or by trying to save data use for highest priority activities. Digital health interventions which depend on data access for functionality (messages, timed reminders, video/photo downloads) may be thwarted with such operational constraints.

Conditional mobile data credits may be an effective approach to improving the uptake of digital interventions because they directly target this barrier to engagement^15^. Although mobile credits have been effective in increasing offline health behaviors in LMICs,^16–18^ limited research examines how mobile data credits can maximize the duration of user engagement with digital health interventions. Understanding how to increase engagement in these platforms may be particularly relevant for interventions that are time-based, such as vaccine reminders, annual health screenings, or coaching on developmental milestones.

The current study, conducted in high-altitude Andean Peru, examines whether conditional mobile data credits increased the duration and frequency of caregivers’ engagement with *Afini,* an AI-supported ECD intervention. *Afini* provides caregivers with responsive, evidence-based guidance on activities tailored to children’s developmental and health-related needs.^13^ In the parent trial of the current nested RCT, the digital ECD intervention showed positive effects on parenting behaviors and children’s development.^8^ The current study was nested within the digital ECD intervention arm of this larger cluster randomized trial. We re-randomized clusters of families to receive either the digital intervention as usual (control) or the digital intervention with mobile data credits conditional on intervention use (treatment). We hypothesized that conditional data credits would increase both the frequency and duration of individual caregivers’ engagement with the digital intervention and whether increased engagement translated into changes in self-reported parenting views and behaviors or child developmental outcomes, the intended target outcomes of the digital ECD intervention. Finally, to assess the economic value of this strategy,^19^ we also examined implementation costs of this mobile data credit intervention.

## Methods

### Study Design

This study was a nested cluster-randomized controlled trial (c-RCT) evaluating whether conditional mobile data credits increased caregiver engagement with a digital ECD intervention. The trial was nested within a larger three-arm parallel c-RCT conducted in three provinces (San Marcos, Cajabamba, and Cajamarca) in the Cajamarca region of high-altitude Andean Peru.^20^ The parent trial and current nested study were registered at clinicaltrials.gov (NCT05202106 [<u>LINK</u>]) and are closed to new participants. Baseline recruitment took place from Sept 13, 2021 to March 3, 2023 and endline assessments were conducted from Sept 8, 2023, to March 17, 2025. The trial ended upon the completion of endline data collection.

The parent trial enrolled caregiver-child dyads across 164 clusters. Each cluster corresponded to a village in rural areas or to a neighborhood in urban settings, for simplicity of recruitment. Clusters were randomly assigned to one of three groups: control, a biweekly home– visiting intervention, or the digital ECD intervention. The current trial was conducted among the 72 community clusters assigned to the digital intervention group in the parent trial. Among these 72 clusters, 20 clusters were randomly selected to receive the additional mobile data credit intervention; the remaining 52 clusters continued to receive the digital intervention only. Equal clusters were assumed.

The study was approved by the University Peruana Cayetano Heredia institutional review board in Lima, Peru (SIDISI: 202522 and 210652) and the Ethics Commission for Northwest and Central Switzerland (EKNZ: AO_2021-00002). All participants signed an informed consent at time of enrollment, after cluster randomization. No important changes were made to the trial after it commenced.

### Participants

Participants were caregivers of children 3-9 months at baseline who had been enrolled in the digital ECD intervention arm of the parent trial.^8^ Caregivers were recruited by data collectors through door-to-door visits. Descriptive characteristics were recorded at baseline, including sociodemographic characteristics such as caregiver sex, age, and education, as well as digital access measures such as smartphone ownership, use of social media, access to household internet signal inside the home, and smartphone as the primary source of internet.

All participants within the intervention arm of the parent trial (N = 1,174, Female = 1,151) were assessed for eligibility for the current nested mobile data credit trial. Eligibility for this trial required possessing a phone number that would accept pay-as-you-go top ups through either *Claro* or *Bitel,* the two regional mobile providers through which small digital data credit transfers could be most affordably administered.

### Randomization and Masking

The parent trial used stratified constrained randomization to allocate community clusters to the three parent trial groups. Full details are reported in the study protocol.^8,20^ For the nested mobile data credit trial, 20 of the 72 clusters were assigned to treatment by G.F., selected via a simple random draw in Stata, stratified by province.^22^ This ratio was selected for costs of administering the mobile data credits and to not greatly sway parent trial results. All eligible caregivers within these 20 clusters (147 caregivers) were assigned to receive mobile data credits in addition to the standard digital ECD intervention. 3 control clusters had no eligible caregivers; as such, the remaining 49 clusters (333 caregivers) received the digital ECD intervention only.

Because the mobile data credit intervention involved visible transfers to participants’ phones and related reminder messages, treatment assignment could not be concealed or blinded. Study staff conducting endline child assessments were not informed of random allocation, although full masking could not be guaranteed in the field. Data analysts were not masked to treatment assignment.

### Procedures

At enrollment, each participant (treatment and control) received access to the *Afini* platform, a multichannel, AI-supported digital intervention designed to assist caregivers in healthy and developmentally appropriate child-rearing practices.^8^ Data collectors introduced caregivers to the platform, supported the setup on their phones, and briefly trained them in its use.^20^ This intervention enabled conversations between participants and an AI-supported virtual tutor that could assess children’s developmental stages and provide relevant recommendations and activities. The system used an AI-supported recommendation engine to select expert-generated content from a pre-approved library based on caregiver engagement and reported feedback. The AI system was used for content recommendation only and did not generate or modify intervention content. Further details on the *Afini* system are provided in the parent trial report and its Supplementary Materials. The platform was available through a mobile application, Facebook Messenger, and WhatsApp. During this same visit, caregivers answered a baseline survey on sociodemographic and household characteristics, caregiving practices, child health, and child development.

Caregivers assigned to the mobile data credit treatment received information about the conditional mobile data credits from data collectors. Caregivers were eligible to receive small mobile data credits every two weeks from enrollment until the end of the intervention period. Credits were conditional on engaging with the digital ECD intervention (e.g., sent or read a message, clicked on a link) at least once during the corresponding two-week period. Mobile data credits were transferred directly to participants’ mobile phones, via third-party services *Reloadly* and *DTOne*. Mobile data credits were valued at approximately 3 Peruvian soles (0.87 USD), corresponding to the smallest data package available. This typically provided about 250 MB of data and, depending on provider, 2-4 weeks of unlimited calling and WhatsApp or Facebook Messenger texting. The exact benefits shifted during the trial due to changes in mobile carrier and data aggregator policies. Participants in the treatment group received an automated reminder if they were at risk of missing a mobile data credit during a two-week period. After a participant failed to engage for two consecutive two-week periods, reminder messages and credits stopped.

Caregivers in the comparison group received the standard digital ECD intervention only, including regular encouragement to use the system, but received no information about or access to mobile data credits. All participants—treatment and control—received one in-person endline data collection visit when children were on average 29 months old, during which secondary outcomes were assessed.

### Outcomes

The primary outcomes pertain to individual caregivers’ frequency and duration of engagement with the digital ECD intervention, as well as group-wide retention (proportion of sample actively using the ECD intervention at defined follow-up intervals). For all engagement outcomes, engagement was defined as any recorded log-in or interaction with the digital ECD intervention within the two-week period between mobile data credit allocations. Engagement outcomes were calculated from platform log data over the first 82 weeks after each caregiver registered on the digital platform. This observation window was selected because 82 weeks was the minimum duration of log data available for all participants, allowing engagement outcomes to be calculated over a common follow-up period for the full analytic sample. Caregivers with no recorded platform activity were coded as having 0 active weeks and 0 weeks of engagement duration.

*Frequency* of engagement was defined as the total number of weeks, from week 1 to week 82 after registration, in which the caregiver had any recorded activity on the platform. This outcome was analyzed as a continuous count variable, with possible values ranging from 0 to 82 active weeks. Next, *duration* was calculated as the number of weeks between first and last recorded interaction. Last, *group-wide retention* examined the proportion of active individual users of the digital intervention at prespecified follow-up intervals. Caregivers were coded as retained at 3, 6, 9, and 12 months if they had any recorded platform activity within ±2 week of 13, 26, 39, and 52 weeks after registration, respectively. Each retention outcome was analyzed as a binary indicator and summarized as the proportion of caregivers active at each interval.

Secondary outcomes were parenting views and behaviors and child development. Parenting views and behaviors were assessed at endline using the Nurturing Parenting Beliefs and Behaviors Scale (NPBBS)^23^, a locally developed measure with validity evidence supporting its cultural relevance. The measure includes a 15-item views (beliefs, attitudes, and knowledge) subscale (Cronbach’s α=0.83) and an 11-item behaviors index. Caregivers responded to each item using a 3-point Likert scale (e.g., not at all (*nada*), a little (*poco*), or a lot (*mucho*). An example item for the views subscale includes, “In your opinion, is it worth reading books or telling stories to young children before they learn to speak?”; an example item from the behaviors subscale includes “How often do you ask (child’s name) to use words to express what he/she needs or wants?” Scores were calculated by averaging items within subscales, with higher scores representing more nurturing parenting views and behaviors, respectively. Child development was also assessed at endline, using the long form of the Global Scales for Early Development (GSED), a direct assessment for children younger than 3 years developed for culturally diverse, low-resource settings.^24^ The assessment uses an age– and ability-adaptive subset of 155 items to capture developmental milestones across domains and produces an overall D-score.

Because the intervention involved low-value mobile data credits and access to digital parenting content, no trial-specific harms were expected and adverse events were not prespecified as formal outcomes. No adverse events related to the trial were reported during the study period.

### Sociodemographic and Developmental Covariates

The primary intention-to-treat (ITT) models were unadjusted. As a sensitivity analysis, we repeated the main models using the pre-specified individual covariates set from the parent trial. Child covariates included sex (1= female, 0 = male); low birthweight (1=birthweight below 2.5 kg, 0 = at or above 2.5 kg), age in months, baseline child development, and baseline home stimulation. Baseline child development was measured using the Caregiver Reported Early Development Instruments (CREDI), a caregiver-reported, population-level measure of ECD designed for children from birth to age 3 years and intended to capture overall developmental status.^25^ Home stimulation was measured using six items adapted from the Multiple Indicator Cluster Survey’s Family Care Indicators, asking whether an adult engaged with the child in the past 3 days in activities such as reading or looking at books or taking the child outside.^26^ Caregiver covariates included mother’s and father’s age (in years), and categorical indicators for maternal and paternal education: minimal education (reference group), completed primary education, completed secondary education, and some tertiary education. Household covariates included father presence in the household (1=yes, 0=no), number of children, number of adults, and household socioeconomic status, measured using an asset-based index generated by principal components analysis and divided into quintiles based on the first principal component in the parent trial.^26^

### Statistical Analysis

We summarized baseline sociodemographic characteristics, community connectivity, and individual phone-use measures for the analytic sample overall and by mobile data credit assignment. We estimated the effect of mobile data credits on caregiver engagement, parenting outcomes, and ECD using an ITT approach among the 480 caregivers in the nested trial. For the primary engagement outcomes, an absence of recorded activity during a two-week period was coded as no engagement. For secondary outcomes assessed at endline, analyses excluded participants without observed outcome data. For continuous outcomes, we used linear regression models with randomized assignment to mobile data credits as the main predictor. Separate models were estimated for each engagement, parenting, and ECD outcome. For binary outcomes, we analyzed linear probability models. We used standard errors cluster-adjusted at the community-cluster level for all models, reflecting the unit of randomization.

Primary models were unadjusted. In sensitivity analyses, we repeated these analyses, using the prespecified covariates (sociodemographic or developmental characteristics of the child, caregiver, and household) from the parent trial to maintain consistency.^8^ Models examining the GSED used the raw D-score and adjusted for the child’s age at assessment. For descriptive visualization, we used platform log-in data from the first 52 weeks after registration to calculate the proportion of caregivers with any recorded activity in each week, separately by randomized assignment group. We plotted locally weighted smoothed curves with pointwise 95% CIs to visualize weekly active use over the first year.

Finally, to assess implementation fidelity and provide perspectives on intervention costs, we reported descriptive statistics (mean, minimum, maximum, standard deviation) for the number of mobile data credits received by participants, the proportion credits successfully fulfilled, and the costs of mobile data credits per participant and per use. No interim analyses were conducted, and no stopping guidelines were specified. Analyses were conducted using STATA.^22^

## Results

Figure 1 shows the trial profile CONSORT flow diagram for the nested trial.^27^ Participants were recruited between September 13, 2021 to March 3, 2023. Of the 1174 caregivers assigned to receive the digital ECD intervention in the parent trial, 480 caregivers had eligible phone providers and data packages (pay-as-you-go) and were included in the analytic sample, and 694 were ineligible. Supplementary Table S1 compares caregivers with eligible and ineligible phone plans in the parent digital intervention cohort. 147 caregivers within 20 clusters were assigned to mobile data credits, and 333 caregivers within 52 clusters were assigned to the digital intervention only. As disengagement was the primary outcome, no participants (N = 480; 147 mobile data credit, 333 control) were considered to have dropped out of the study. Endline participation rates did not differ between groups, with 83.0% (n=122/147) treatment vs 85.3% (n=284/333) control, difference –2.3 pp, 95% CI –8.1 to 3.5 pp, p=0.430.

**Fig 1.**
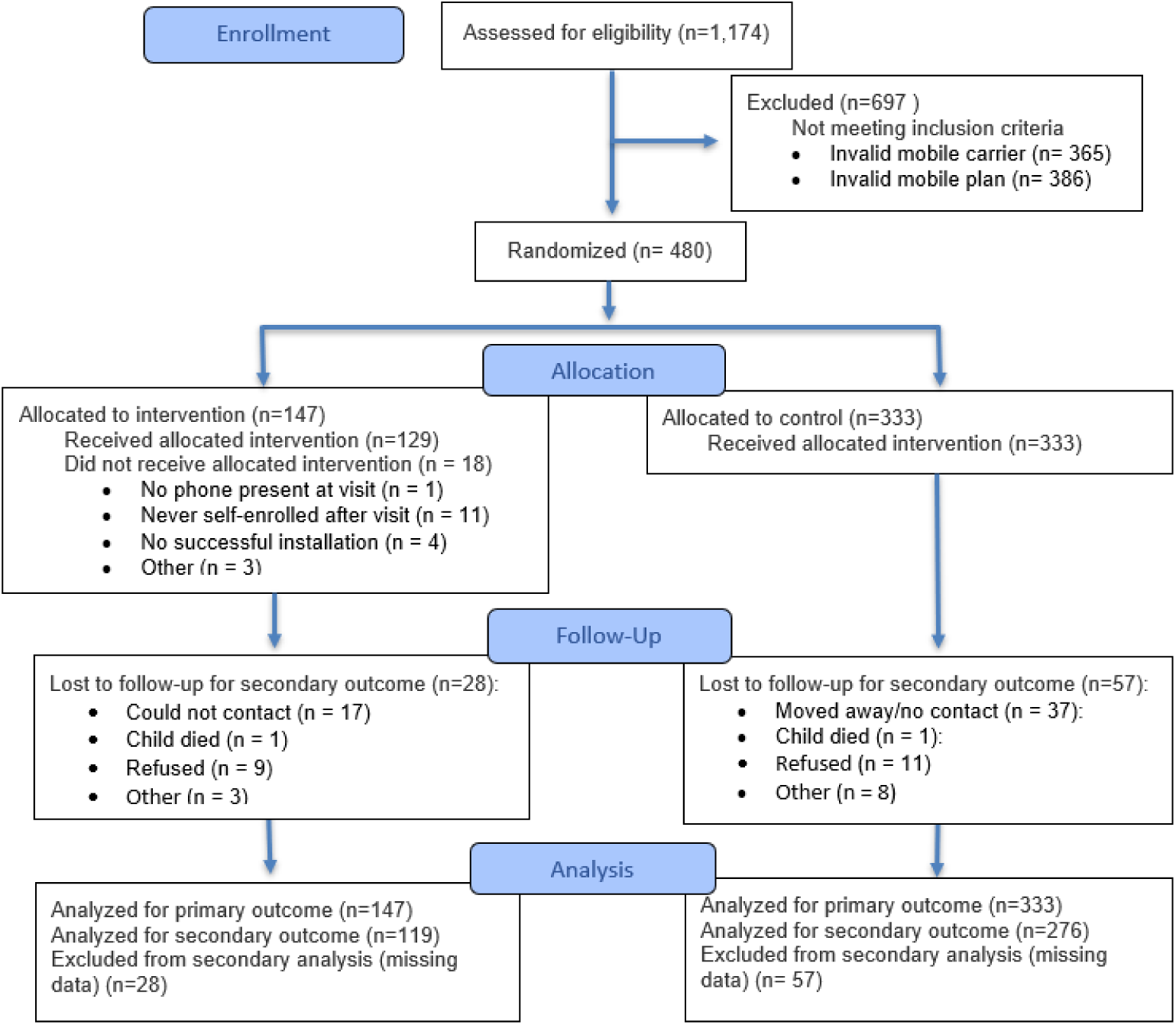
CONSORT Flow Diagram.

Baseline characteristics of the analytic sample are shown in Table 1, both as totals and stratified by assignment to treatment and control groups. Children in the sample were 48% (229/480) female, 50% (234/473) of caregivers had completed basic education, and 90% (434/480) of households had fathers present. Most households had internet signal inside the home (89%; 425/476), and 90% (406/450) of caregivers reported that a smartphone was their main source of internet access. Denominators varied between 450 and 480 because of item-level non-response.

**Table 1:** Demographics of analytic sample.

| Variable | Total Analytic Sample |  | Control Group |  | Mobile data credit group |  |
| --- | --- | --- | --- | --- | --- | --- |
|  | % or Mean | n or SD | % or Mean | n or SD | % or Mean | n or SD |
| Total |  | 480 |  | 333 |  | 147 |
| **Child Characteristics** |  |  |  |  |  |  |
| Child is female | 48% | 229 | 48% | 159 | 48% | 70 |
| Child birth weight <2.5 kg | 7% | 33 | 7% | 24 | 6% | 9 |
| Child age in months | 4.99 | 1.88 | 5.00 | 1.91 | 4.98 | 1.83 |
| Overall child development at baseline (CREDI) | 0.15 | 0.59 | 0.13 | 0.60 | 0.20 | 0.56 |
| Child stimulation activities | 3.43 | 1.20 | 3.43 | 1.20 | 3.43 | 1.20 |
| **Caregiver Characteristics** |  |  |  |  |  |  |
| Mother age | 26.64 | 6.75 | 26.65 | 6.73 | 26.62 | 6.80 |
| Mother education |  |  |  |  |  |  |
| Minimal education | 12% | 55 | 12% | 39 | 11% | 16 |
| Completed primary education | 20% | 96 | 20% | 67 | 20% | 29 |
| Any secondary education | 49% | 234 | 49% | 160 | 51% | 74 |
| Any tertiary education | 19% | 88 | 19% | 62 | 18% | 26 |
| Father age | 30.39 | 7.76 | 30.20 | 7.85 | 30.83 | 7.58 |
| Father education |  |  |  |  |  |  |
| Minimal education | 5% | 22 | 5% | 15 | 5% | 7 |
| Any primary education | 13% | 56 | 14% | 41 | 12% | 15 |
| Any secondary education | 56% | 235 | 55% | 159 | 59% | 76 |
| Some tertiary education | 25% | 104 | 25% | 73 | 24% | 31 |
| **Household Characteristics** |  |  |  |  |  |  |
| Father is present | 90% | 434 | 90% | 301 | 90% | 133 |
| Number of children in household | 0.96 | 1.11 | 0.93 | 1.14 | 1.03 | 1.03 |
| Number of adults in household | 2.44 | 1.02 | 2.47 | 1.07 | 2.35 | 0.90 |
| SES index | 2.96 | 1.37 | 2.92 | 1.35 | 3.07 | 1.41 |
| Urban community | 34% | 164 | 38% | 127 | 25% | 37 |
| Agriculture is main income | 48% | 229 | 49% | 163 | 45% | 66 |
| **Connectivity measured in community center** |  |  |  |  |  |  |
| Number of phone providers in community (out of 4) | 3.53 | 0.67 | 3.48 | 0.68 | 3.64 | 0.64 |
| Community receives signal from eligible carriers | 99% | 474 | 99% | 331 | 97% | 143 |
| Installation link opens using eligible carriers | 87% | 417 | 90% | 301 | 79% | 116 |
| **Individual Phone Use** |  |  |  |  |  |  |
| Caregiver owns personal smartphone | 83% | 396 | 83% | 274 | 83% | 122 |
| Caregiver uses FB Messenger | 75% | 360 | 76% | 250 | 75% | 110 |
| Caregiver uses WhatsApp | 90% | 429 | 90% | 297 | 90% | 132 |
| Household receives internet signal inside home | 89% | 425 | 90% | 299 | 88% | 126 |
| Smartphone is main source of internet | 90% | 406 | 90% | 285 | 90% | 121 |
| Daily internet use (minutes) | 70.47 | 99.29 | 70.17 | 102.47 | 71.16 | 92.02 |
| *Phone operator* |  |  |  |  |  |  |
| Claro | 52% | 248 | 47% | 158 | 61% | 90 |
| Bitel | 48% | 232 | 53% | 175 | 39% | 57 |
| Successfully completed installation at baseline visit | 79% | 379 | 79% | 263 | 79% | 116 |
| Endline participation | 85% | 406 | 85% | 284 | 83% | 122 |

*Note.* Mobile data credit group assignment was randomized at the neighborhood-cluster level. Denominators vary because of missing data.

Unadjusted ITT estimates for primary outcomes are shown in Table 2. Mobile data credits also increased engagement frequency. Over the 82-week observation window, caregivers in the mobile data credit group were active for a mean of 26.2 weeks, compared with 19.7 weeks in the control group (95% CI = 1.56 to 11.44 weeks, p=0.011). Average engagement duration (time between first and last use) was also longer in the mobile data credit group than in the control group, but this difference was not statistically significant (51.0 weeks vs 44.8 weeks, 95% CI = –1.03 to 13.82 weeks, p=0.102). The intracluster correlation coefficient for engagement frequency was 0.005 (mean cluster size = 7.0).

**Table 2:**
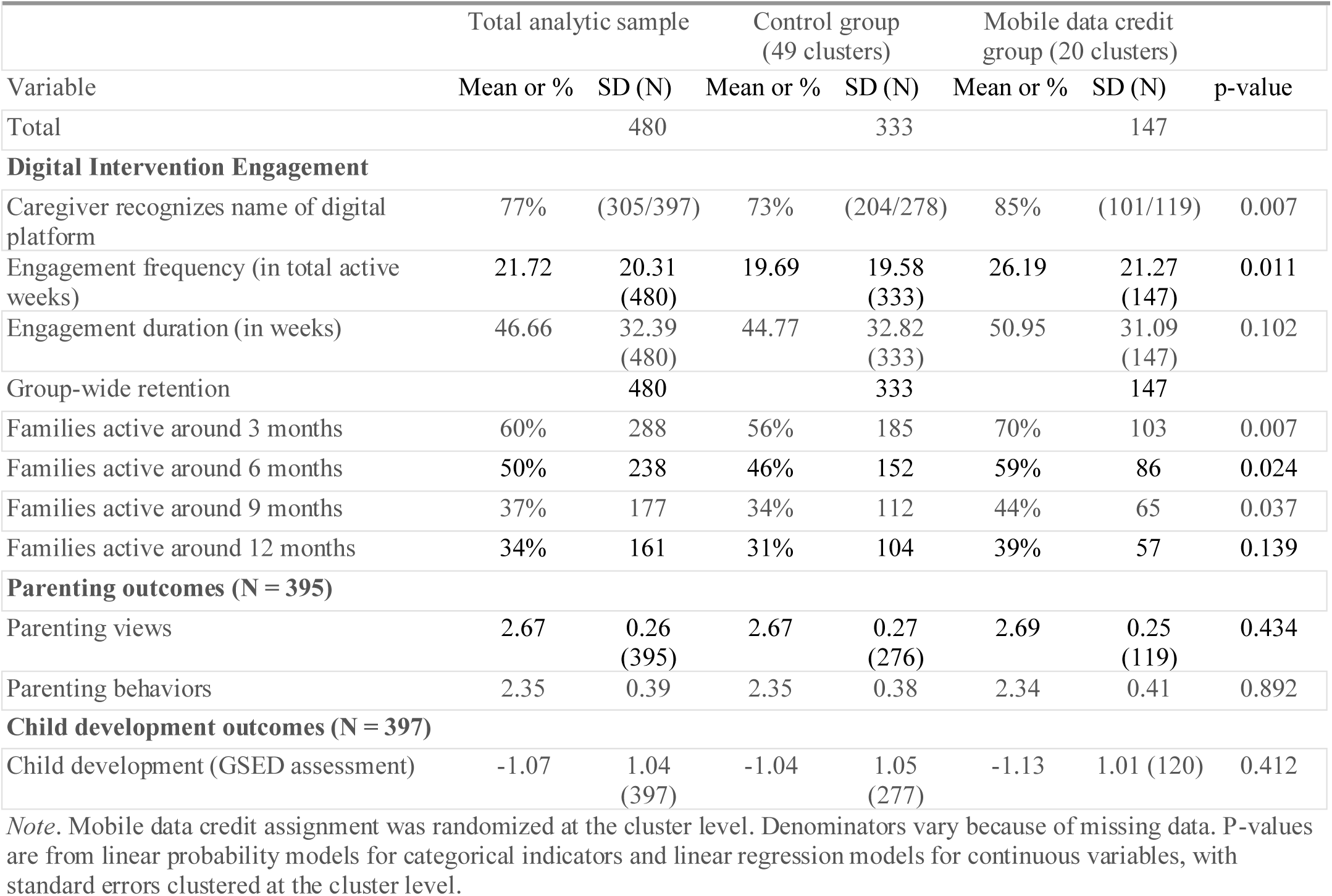
Primary and secondary outcomes by mobile data credit assignment.

| Variable | Total analytic sample |  | Control group<br>(49 clusters) |  | Mobile data credit<br>group (20 clusters) |  | p-value |
| --- | --- | --- | --- | --- | --- | --- | --- |
|  | Mean or % | SD (N) | Mean or % | SD (N) | Mean or % | SD (N) |  |
| Total |  | 480 |  | 333 |  | 147 |  |
| **Digital Intervention Engagement** |  |  |  |  |  |  |  |
| Caregiver recognizes name of digital platform | 77% | (305/397) | 73% | (204/278) | 85% | (101/119) | 0.007 |
| Engagement frequency (in total active weeks) | 21.72 | 20.31<br>(480) | 19.69 | 19.58<br>(333) | 26.19 | 21.27<br>(147) | 0.011 |
| Engagement duration (in weeks) | 46.66 | 32.39<br>(480) | 44.77 | 32.82<br>(333) | 50.95 | 31.09<br>(147) | 0.102 |
| Group-wide retention |  | 480 |  | 333 |  | 147 |  |
| Families active around 3 months | 60% | 288 | 56% | 185 | 70% | 103 | 0.007 |
| Families active around 6 months | 50% | 238 | 46% | 152 | 59% | 86 | 0.024 |
| Families active around 9 months | 37% | 177 | 34% | 112 | 44% | 65 | 0.037 |
| Families active around 12 months | 34% | 161 | 31% | 104 | 39% | 57 | 0.139 |
| **Parenting outcomes (N = 395)** |  |  |  |  |  |  |  |
| Parenting views | 2.67 | 0.26<br>(395) | 2.67 | 0.27<br>(276) | 2.69 | 0.25<br>(119) | 0.434 |
| Parenting behaviors | 2.35 | 0.39 | 2.35 | 0.38 | 2.34 | 0.41 | 0.892 |
| **Child development outcomes (N = 397)** |  |  |  |  |  |  |  |
| Child development (GSED assessment) | -1.07 | 1.04<br>(397) | -1.04 | 1.05<br>(277) | -1.13 | 1.01 (120) | 0.412 |

*Note.* Mobile data credit assignment was randomized at the cluster level. Denominators vary because of missing data. P-values are from linear probability models for categorical indicators and linear regression models for continuous variables, with standard errors clustered at the cluster level.

The proportion of families active at each interval was higher in the mobile data credit group than in the control group at 3 months (70% vs 56%, 95% CI = 4.7 to 25.0 pp, p=0.007), 6 months (59% vs 46%, 95% CI = 2.0 to 24.3 pp, p=0.024), and 9 months (44% vs 34%, 95% CI = 0.6 to 20.5 pp, p=0.037). At 12 months, retention remained numerically higher in the mobile data credit group, but the difference was not statistically significant (39% vs 31%, 95% CI = –2.6 to 17.6 pp, p=0.139). Figure 2 illustrates these patterns, showing weekly active use by mobile data credit group over the first 12 months, with locally weighted smoothed trends and 95% CIs.

**Fig 2.**
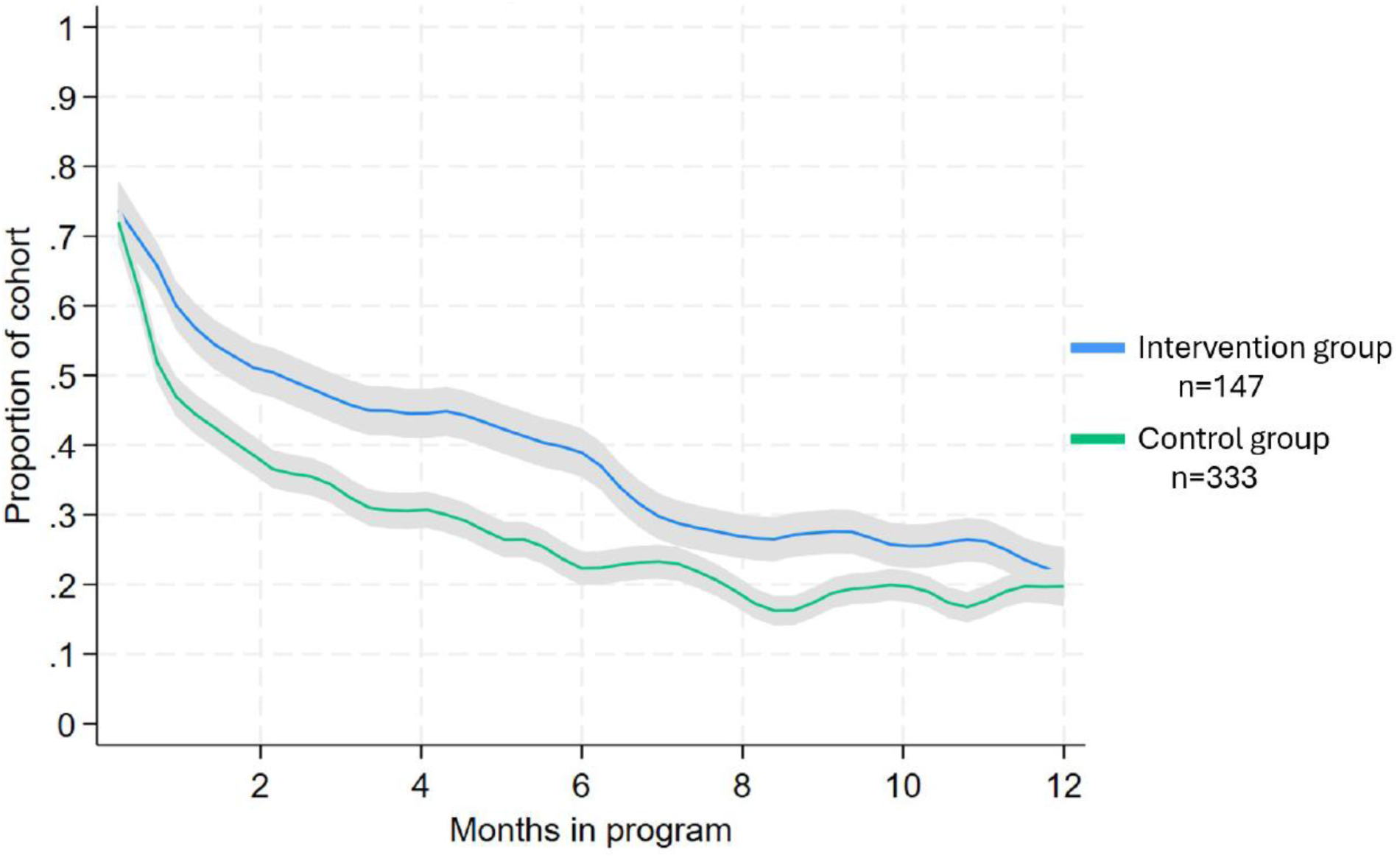
Weekly Active Use by Treatment Status.

Secondary outcomes are also shown in Table 2. We found no evidence that mobile data credits improved caregivers’ self-reported views or behaviors or ECD outcomes at endline. Specifically, mean parenting views scores were similar in the mobile data credit and control groups (2.69 vs 2.67, 95% CI = –0.039 to 0.089, p=0.434), as were parenting behavior scores (2.34 vs 2.35, 95% CI = –0.075 to 0.065, p=0.892). Age-adjusted child development scores at endline also did not differ significantly between groups (–1.13 vs –1.04, 95% CI = –0.300 to 0.124, p=0.412). Adjusted sensitivity ITT analyses were consistent with the primary findings and are presented in Supplemental Table S2.

Implementation fidelity and cost descriptives are presented in Table 3. Participants were intended to receive mobile data credits worth approximately 3 Soles (approximately 0.87 USD) every 2 weeks that they actively engaged with the digital intervention. Due to changing fees charged by the mobile carriers and credit aggregators to administer the credits, we spent between 0.99 USD and 2.56 USD (mean 1.29 USD, SD 0.41, 95% CI 1.27 to 1.31) per active recipient every 2 weeks. This range still covered our intended minimum, providing data to interact with the digital intervention at least one day every 15 days. These administrative costs equate to 18.23 USD, SD 10.12 (median 18.06, range 2.58-32.25, 95% CI 16.36 to 20.11, n=112 recipients) over the course of one year. Based on our impact findings, we equate this to an average of 2.80 USD per additional week of active participation, on average. Mobile data credits were successfully sent to participants 94.7% of the time; barriers to delivery included turnover in aggregator companies (which were often start-ups) and technical glitches with data delivery. Among participants in the mobile data credit group who responded to questions about usefulness, 58.6% reported that credits helped “very much”, 40.0% “a little”, 1.4% “did not help at all” (*N=*70).

**Table 3:**
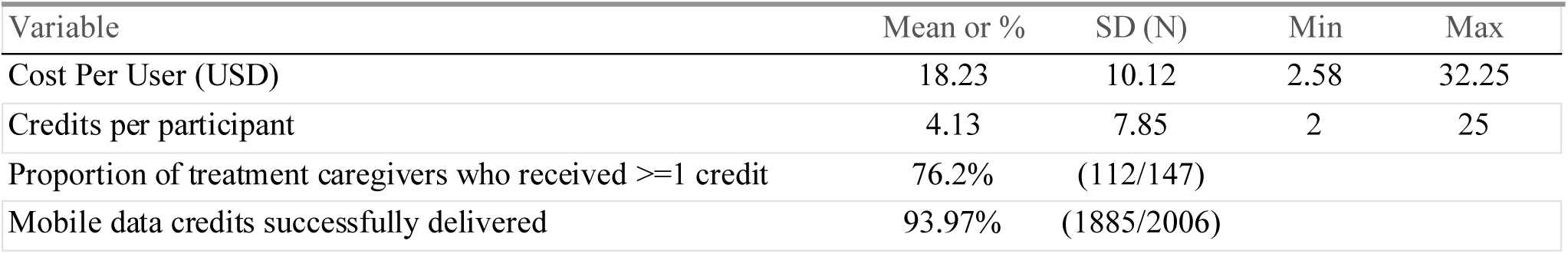
Implementation and cost descriptives.

## Discussion

Conditional mobile data credits modestly extended caregivers’ engagement with an AI-supported digital intervention in a low-resource population in rural, high Andean Peru. Caregivers assigned to receive mobile data credits conditional on intervention use remained engaged with the intervention for nearly 7 additional weeks, on average, compared to the average 20 weeks of engagement by caregivers in the control group. We also found that a higher percentage of the treatment group was active in the digital ECD intervention at 3, 6, and 9 months, compared to the control group. These average gains aligned with our hypothesis that small, conditional data transfers can improve sustained use of digital general prevention interventions in low-resource settings. This likely functions by reducing the perceived expense of engagement, which was supported by caregivers reporting the credit was useful. This intervention may also function as a behavioral boost by supporting participants’ decision making towards intended longer-term rewards such as healthy child development.^28^

Contrary to our hypotheses, we did not find effects of conditional mobile data credits on secondary outcomes (parenting views, behaviors, and child development). This may be partly due to intervention design, as caregivers had access to all aspects of the digital intervention upon enrollment and may experience diminishing value to engagement over time; it could also be the case that null effects were a result of imprecision from a small sample size, or from fadeout of effects (secondary outcomes were typically measured 40 weeks after impacts of the incentives ceased).^29^

The total cost of providing mobile data credits to each participant was on average 18.23 USD over the course of one year. The broader digital ECD intervention was estimated to cost 41.4 USD per child, with an estimated cost of 414 USD per standard deviation increase in child development. Thus, while the expenses described are relatively small, the added value of mobile data credit incentives likely depends on the design and aims of a digital health intervention.^19^ The benefits of extending engagement may be most useful for digital interventions that require frequent contact over time, such as supports with vaccination, medication adherence, or antenatal care. It may also be more valuable for interventions designed to shift collective practices or community norms, as past research has shown that mobile data credits can improve group-wide retention, with plausible positive spillover effects on developmentally supportive parenting behaviors within the community.^30^

Some limitations should be considered when interpreting the current findings. First, this study focused on incentives for engagement in a multichannel ECD platform in high-altitude Andean Peru, and may not generalize to other contexts due to differences in individuals’ digital access and literacy, family’s financial pressures, data carriers’ technical capacity for conditional mobile data credits, or differences in the community’s perceptions of the intervention.^16^ Future research should examine the impacts of this approach in different communities and with digital health interventions that depend on time-based mechanisms to achieve success. Next, implementation logistics shaped our study in meaningful ways. Two of four mobile carrier providers supported affordable automated data transfers and could only support data credits to individuals without automatically renewing data contracts. Beyond reducing participant eligibility, (*N* parent cohort = 1174, *N* eligible = 480), this limits the population to whom these findings generalize, as we found sociodemographic differences between individuals with eligible and non-eligible phone providers. Practically, differences in eligibility is also an equity concern: digital incentive systems may systematically exclude participants whose providers or devices are incompatible with automated delivery.

Relatedly, prices to administer these incentives fluctuated (typically increased) over the duration of the study, and mobile carriers and aggregators provided different levels of transparency as to whether incentives were successfully delivered. These practical limitations shape the feasibility of implementing this approach in novel contexts. Finally, while the current study aimed to support participants’ agency by tying incentives to the use of a strengths-based, responsive intervention, caregivers’ perceptions of pressure to participate due to incentives was not explored. Future research should explore perceptions of and technical capacity for this approach across regions.

## Conclusion

The current study examined how conditional mobile data credits shaped the frequency and duration of engagement with an AI-supported early childhood digital health platform designed to support parenting processes and child development. Conditional mobile data credits significantly improved engagement with the digital intervention, but these improvements did not translate into significant effects on caregivers’ childrearing views or behaviors or child outcomes 40 weeks later. These findings suggest that mobile data credits can be a low-cost strategy for improving engagement with digital interventions. Whether or not these incentives are cost-effective will depend on the health or developmental benefits that can be achieved through additional use. Researchers, practitioners, and policymakers interested in utilizing mobile data credit incentives should consider relative cost, the equity of digital infrastructure, and the extent to which sustained engagement is expected to translate into meaningful health or developmental gains.

## Data Availability

Data will be shared for verification purposes on request. According to the permissions granted in our ethical review, the dataset and data will be provided (fully anonymised) for verification purposes on request only.

## Acknowledgments

We thank the fieldworkers who recruited participants and collected data for this trial, and the data aggregators who administered top-ups.

## References

1. GSMA. The Mobile Economy 2023. GSM Association, https://www.gsma.com/solutions-and-impact/connectivity-for-good/mobile-economy/wp-content/uploads/2023/03/270223-The-Mobile-Economy-2023.pdf (2023).

2. UNESCO. Literacy Rates Continue to Rise from One Generation to the Next. Fact Sheet 45, UNESCO Institute for Statistics, https://uis.unesco.org/sites/default/files/documents/fs45-literacy-rates-continue-rise-generation-to-next-en-2017_0.pdf#page=2.18 (September 2017, accessed 15 September 2025).

3. Clark H, Coll-Seck AM, Banerjee A, et al. A future for the world’s children? A WHO–UNICEF–Lancet Commission. The Lancet 2020; 395: 605–658.

4. Geldsetzer P, Flores S, Wang G, et al. A systematic review of healthcare provider-targeted mobile applications for non-communicable diseases in low– and middle-income countries. Npj Digit Med 2022; 5: 99.

5. Jäggi L, Hartinger SM, Fink G, et al. Parenting in the Digital Age: A Scoping Review of Digital Early Childhood Parenting Interventions in Low– and Middle-Income Countries (LMIC). Public Health Rev 2025; 45: 1607651.

6. Black MM, Walker SP, Fernald LCH, et al. Early childhood development coming of age: science through the life course. The Lancet 2017; 389: 77–90.

7. Head KJ, Noar SM, Iannarino NT, et al. Efficacy of text messaging-based interventions for health promotion: A meta-analysis. Soc Sci Med 2013; 97: 41–48.

8. Fink G, McCoy DC, Jäggi L, et al. Digital support systems to improve child development in Peru: A cluster-randomized controlled open-label trial. Sci Adv 2026; 12: eaeb9403.

9. Balsa AI, Gómez Muzzio E, González ML, et al. Crianza Positiva: Combining Group Workshops and E-Messages to Strengthen Parenting Competences. Child Youth Care Forum 2024; 53: 719–740.

10. Janowski R, Cluver LD, Shenderovich Y, et al. Optimizing Engagement With a Smartphone App to Prevent Violence Against Adolescents: Results From a Cluster Randomized Factorial Trial in Tanzania. J Med Internet Res 2025; 27: e60102.

11. Awah I, Green O, Baerecke L, et al. ‘It provides practical tips, practical solutions!’: acceptability, usability, and satisfaction of a digital parenting intervention across African countries. Psychol Health Med 2022; 27: 107–123.

12. Dadds MR, Sicouri G, Piotrowska PJ, et al. Keeping Parents Involved: Predicting Attrition in a Self-Directed, Online Program for Childhood Conduct Problems. J Clin Child Adolesc Psychol 2019; 48: 881–893.

12. Castellanos A. UNICEF Innovation Fund Graduate: Afinidata | UNICEF Office of Innovation. *UNICEF Office of Innovation*, https://www.unicef.org/innovation/FundGraduate/Afinidata (2020, accessed 17 June 2026).

14. Jäggi L, Aguilar L, Alvarado Llatance M, et al. Digital tools to improve parenting behaviour in low-income settings: a mixed-methods feasibility study. Arch Dis Child 2023; 108: 433–439.

15. Chandrasekar A, Warren E, Free C, et al. mHealth interventions for postpartum family planning in LMICs: A realist review. PLOS Glob Public Health 2024; 4: e0003432.

16. Saunders MJ, Pereboom M, Alvarez JL, et al. Incentives in immunisation campaigns in low– and middle-income countries: a scoping review mapping evidence on effectiveness and unintended consequences. BMJ Glob Health 2025; 10: e019662.

17. Banerjee A, Chandrasekhar A, Duflo E, et al. Evaluating the impact of interventions to improve full immunisation rates in Haryana, India. 2020th edn. International Initiative for Impact Evaluation (3ie). Epub ahead of print September 2020. DOI: 10.23846/TW10IE126.

18. Seth R, Akinboyo I, Chhabra A, et al. Mobile Phone Incentives for Childhood Immunizations in Rural India. Pediatrics 2018; 141: e20173455.

19. Herrick T, Digre P, Yoden SS. Building the evidence base for the economic value of digital health interventions: examples of methods and metrics and opportunities for future directions. Oxf Open Digit Health 2024; 2: ii1–ii3.

20. Hartinger Pena SM, Mäusezahl D, Jäggi L, et al. Digital Support Systems to Improve Child Health and Development in Peru: Protocol for a Randomized Controlled Trial. JMIR Res Protoc 2023; 12: e50371.

21. Moerbeek M. Randomization of Clusters Versus Randomization of Persons Within Clusters: Which Is Preferable? Am Stat 2005; 59: 173–179.

22. StataCorp. 2025. Stata Statistical Software: Release 19. College Station, TX: StataCorp LLC.

23. Hinckley K, Alvarado M, McCoy DC, et al. Development and validation of the Nurturing Parenting Beliefs and Behaviors Scale (NPBBS): Measuring parenting in high Andean Peru. Dev Psychol 2026; 62: 233–253.

24. McCray G, McCoy D, Kariger P, et al. The creation of the Global Scales for Early Development (GSED) for children aged 0–3 years: combining subject matter expert judgements with big data. BMJ Glob Health 2023; 8: e009827.

25. McCoy DC, Waldman M, Fink G. Measuring early childhood development at a global scale: Evidence from the Caregiver-Reported Early Development Instruments. Early Child Res Q 2018; 45: 58–68.

26. Kariger P, Frongillo EA, Engle P, et al. Indicators of Family Care for Development for Use in Multicountry Surveys. J Health Popul Nutr 2013; 30: 472–486.

27. Hopewell S, Chan A-W, Collins GS, et al. CONSORT 2025 statement: updated guideline for reporting randomised trials. BMJ 2025; 389: e081123.

28. Herzog SM, Hertwig R. Boosting: Empowering Citizens with Behavioral Science. Annu Rev Psychol 2025; 76: 851–881.

29. Bailey DH, Duncan GJ, Cunha F, et al. Persistence and Fade-Out of Educational-Intervention Effects: Mechanisms and Potential Solutions. Psychol Sci Public Interest 2020; 21: 55–97.

30. Benjamin-Chung J, Arnold BF, Berger D, et al. Spillover effects in epidemiology: parameters, study designs and methodological considerations. Int J Epidemiol 2018; 47: 332– 347.

